# More rapid, robust and sustainable antibody responses to mRNA COVID-19 vaccine in convalescent COVID-19 individuals

**DOI:** 10.1101/2021.08.04.21261561

**Authors:** Sabrina E. Racine-Brzostek, Jim Yee, Ashley Sukhu, Yuqing Qiu, Sophie Rand, Paul Barone, Ying Hao, He S. Yang, Qing H Meng, Fred S Apple, Yuanyuan Shi, Amy Chadburn, Encouse Golden, Silvia C. Formenti, Melissa M. Cushing, Zhen Zhao

## Abstract

Longitudinal studies are needed to evaluate the SARS-CoV-2 mRNA vaccine antibody response under “real-world” conditions. This longitudinal study investigated the quantity and quality of SARS-CoV-2 antibody response in 846 specimens from 350 subjects: comparing BNT162b2-vaccinated individuals (19 previously diagnosed with COVID-19 [RecoVax]; 49 never been diagnosed [NaïveVax]) to 122 hospitalized unvaccinated (HospNoVax) and 160 outpatient unvaccinated (OutPtNoVax) COVID-19 patients.

NaïveVax experienced a delay in generating SARS-CoV-2 total antibody levels (TAb) and neutralizing antibodies (SNAb) after the 1st vaccine dose (D1), but a rapid increase in antibody levels was observed after the 2^nd^ dose (D2). However, these never reached the robust levels observed in RecoVax. In fact, NaïveVax TAb and SNAb levels decreased 4-weeks post-D2 (p=0.003;p<0.001). For the most part, RecoVax TAb persisted throughout this study, after reaching maximal levels 2-weeks post-D2; but SNAb decreased significantly ∼6-months post-D1 (p=0.002). Although NaïveVax avidity lagged behind that of RecoVax for most of the follow-up periods, NaïveVax did reach similar avidity by ∼6-months post-D1. These data suggest that one vaccine dose elicits maximal antibody response in RecoVax and may be sufficient. Also, despite decreasing levels in TAb and SNAb overtime, long-term avidity maybe a measure worth evaluating and possibly correlating to vaccine efficacy.

## Introduction

As the Coronavirus Disease 2019 (COVID-19) pandemic enters its second year with more than 193 million confirmed cases worldwide,(1) many countries look to effective prophylactic SARS-CoV-2 vaccines to help curb its spread and prevent the thousands of COVID-19 deaths reported daily.(2) In December 2020, the United States Food and Drug Administration issued an emergency use authorization (EUA) for two SARS-CoV-2 mRNA vaccines. Shortly thereafter, New York State commenced their vaccination program by vaccinating healthcare workers.

In addition to clinical trials conducted to determine the safety and efficacy of the mRNA vaccines,(3-5) additional studies began describing the serological response to the vaccines under “real-world” conditions, especially with the onset of the SARS-CoV-2 variants and case reports of vaccine escape.(6-13) Although the initial focus may be on overall antibody levels and differences in the antibody response in previously seropositive versus seronegative vaccine recipients,(6, 8, 11) other humoral antibody response factors need to be considered. Due to their role in inactivating viruses and limiting the number of infected host cells, the presence of neutralizing antibodies are often considered a gold standard in evaluating protective immune responses.(14) Early studies describe differences in the neutralizing response post-vaccination in those previously exposed versus naïve to SARS-CoV-2. (12, 15, 16) Binding avidity, the intrinsic affinity of the antibody-antigen interaction, is another potential factor in evaluating the quality of the antibody response. Studies show that overtime, the low avidity antibodies produced early in the humoral immune response to SARS-CoV-2 mature and strengthen, displaying higher intrinsic affinity.(17-19) However, this has not been studied in SARS-CoV-2 vaccinated individuals.

This study evaluated the dynamics of the antibody response to the BNT162b2 mRNA SARS-CoV-2 vaccine, including total antibody levels (TAb), neutralizing antibody levels and antibody avidity, in 49 non-infected vaccinated (NaïveVax) and 19 previously infected vaccinated (RecoVax) healthcare workers. The vaccine-induced response was then compared to the natural post-infection antibody response in 160 non-vaccinated outpatients with mild COVID-19 symptoms (OutPtNoVax) and 122 non-vaccinated COVID-19 hospitalized acutely infected (HospNoVax) (20) during the early period of the pandemic.

## Results

### Participant Demographics

#### Vaccine study cohorts (prospective)

Participant demographics of the 68 healthcare worker volunteers who had been vaccinated 12/18/2020-2/11/2021 with the BNT162b2 vaccine are summarized in Table 1.

**Table 1.** Demographics of vaccinated cohorts compared to hospitalized COVID-19 patient cohort.

| Characteristic | Vaccinated cohorts |  | Non-Vaccinated cohorts |  |
| --- | --- | --- | --- | --- |
|  | Previously COVID positive (RecoVax) (n=19) | Previously COVID negative (NaïveVax) (n=49) | Hospitalized COVID-19 patients (HospNoVax) (n=122) | Outpatient COVID-19 patients (OutPtNoVax) (n=160) |
| **Age, y** |  |  |  |  |
| **Mean (SD)** | 42.5 (11.6) | 46.3 (13.3) | 65.4 (15.9) | 43.6 (12.0) |
| **Median (IQR)** | 40.6 (32.5 – 55.2)<sup>W</sup> | 45.8 (34.4 – 58.3)<sup>W</sup> | 67.5 (54.0-77.0) | 43.0 (34.0-51.1) |
| **Sex - No. (%)** |  |  |  |  |
| **Female** | 11(58)<sup>F</sup> | 39 (80)<sup>F</sup> | 42 (34) | 110 (69) |
| **Male** | 8 (42)<sup>F</sup> | 10 (20)<sup>F</sup> | 80 (66) | 50 (31) |
| **DAOS to first dose vaccination, days, Median, IQR** | 262 (101.5-275.0) | n/a | n/a | n/a |
| **DAOS to ED visit, days, Median, IQR** | n/a | n/a | 7 (4-10) | n/a |
| **DAOS to outpatient visit, days, Median, IQR** | n/a | n/a | n/a | 47 (43-48) |

Test: W: Wilcoxon rank-sum test p = 0.2693; F: Fisher's exact test p=0.1231
DAOS: days after onset of symptoms

#### RecoVax

19 participants (27.9%), previously diagnosed with symptomatic COVID-19 either by RT-PCR (7/19; 36.8%), prior serology (9/19; 47.4%) or clinically diagnosed with COVID-19 during the early periods of the pandemic when testing was unavailable (3/19; 15.8%). The median time from COVID-19 symptom onset to 1^st^ dose vaccine (D1) in this cohort was 262 days (IQR 102-275). All 19 participants tested positive for SARS-CoV-2 serology when participant sera were evaluated with the Roche Elecsys^®^Anti-SARS-CoV-2 assay, which identifies N protein antibodies produced by infection rather than vaccination since the BNT162b2 vaccine does not include the N-protein.

#### NaïveVax

50 participants, not diagnosed with COVID-19 and did not have antibodies against the N-protein at the onset of the study. However, one participant was excluded due to testing positive for COVID-19 by RT-PCR 6 days after the first vaccine dose. (Figure 1)

**Figure 1.**
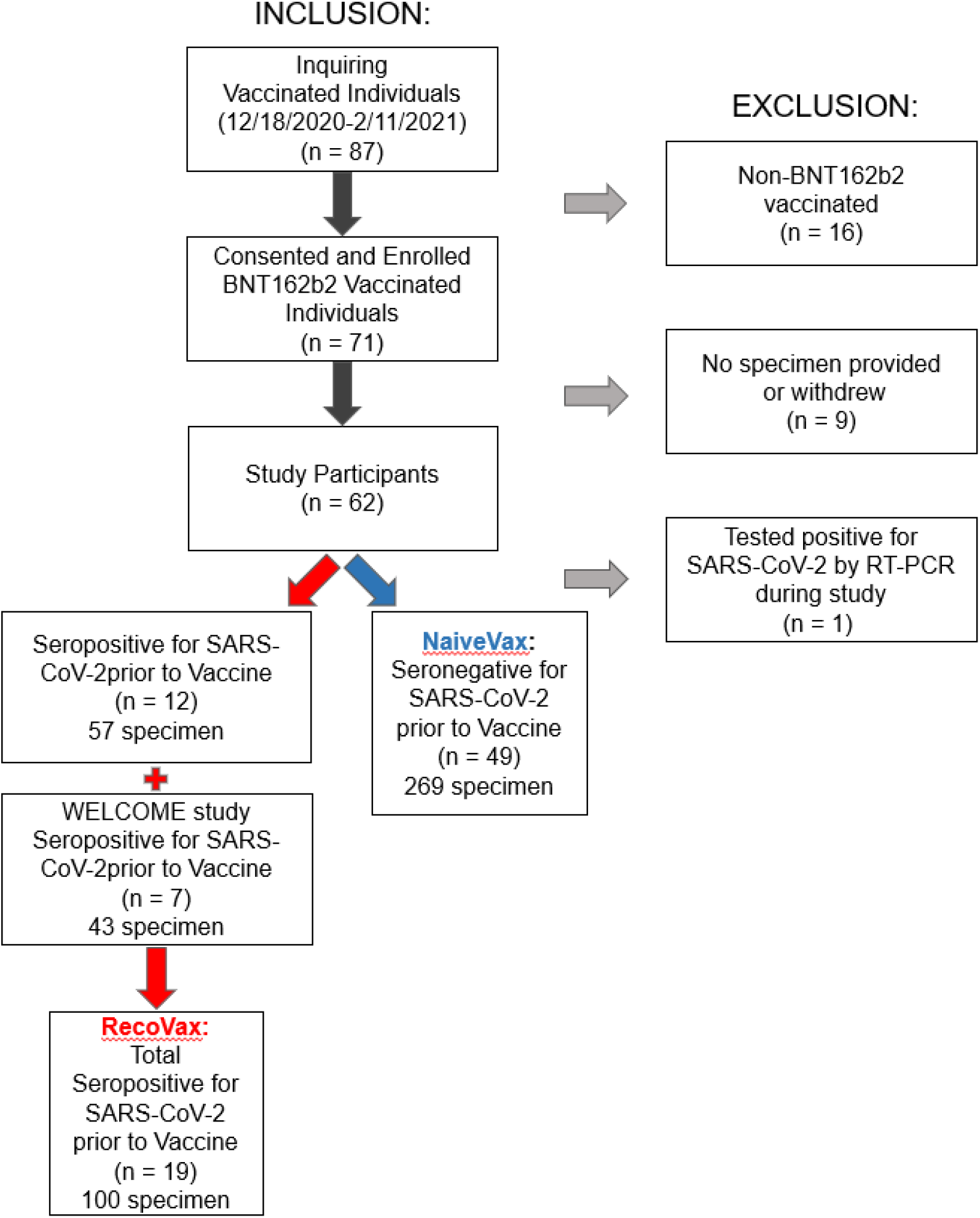
Inclusion and exclusion criteria of the prospective vaccination arm of the study.

### Non-vaccinated cohorts (retrospective)

#### HospNoVax

122 adult patients who had presented to the ED and were subsequently hospitalized at NYP/WCMC during the first month of the pandemic in New York City (3/8/2020-4/7/2020). The antibodies generated by these patients would be most consistent with the initially described SARS-CoV-2 virus. This comparison was prudent as multiple variants have since been described (21) and the vaccine’s design was based on the nucleoside-modified mRNA that encodes the trimerized RBD of the early SARS-CoV-2 spike glycoprotein. (22)

HospNoVax demographics are summarized in Table 1 and further described in a previous validation study.(23) The median time from COVID-19 symptom onset to the ED visit was 7 days (IQR 4-10). In an attempt to estimate time of infection for comparison studies with the vaccinated cohorts, it was estimated that the time of infection was 5 days prior to date of symptom onset.(24, 25) Therefore, the median estimated time of infection to the ED visit was 12 days (IQR 7-15). Also of note, 39/122 (32%) were intubated, and 32/122 (26.2%) died during their hospitalization.(23)

#### OutPtNoVax

160 adult patients who had presented to an outpatient clinic in person or via video visit and had been tested for suspicion of prior COVID-19 infection. Specimen for SARS-CoV-2 serology testing occurred during the time period 4/30/2020 -5/20/2020. As with the hospitalized patients, antibodies generated by these patients would be most consistent with the initially described SARS-CoV-2 virus. This outpatient cohort’s demographics are summarized in Table 1. The median time from COVID-19 symptom onset to the outpatient visit was 47 days (IQR 43-48). As with the hospitalized cohort, it was estimated that the time of infection was 5 days prior to date of symptom onset. Therefore, the median estimated time of infection to the outpatient visit was 52 days (IQR 48-53).

### Quantitative antibody response during the first 2 months post-vaccination compared to post-infection

Using regression models, it was found that HospNoVax had a gradual rise in anti-SARS-CoV-2 TAb antibody levels up to 33 days post-infection with the TAb RFU increasing 277 RFU/day (p<0.001), at which point the levels began to plateau and no significant change was observed to the last follow-up period of 61 days (mean 7003 RFU, coefficient 23 p=0.735). (Figure 2A)

**Figure 2.**
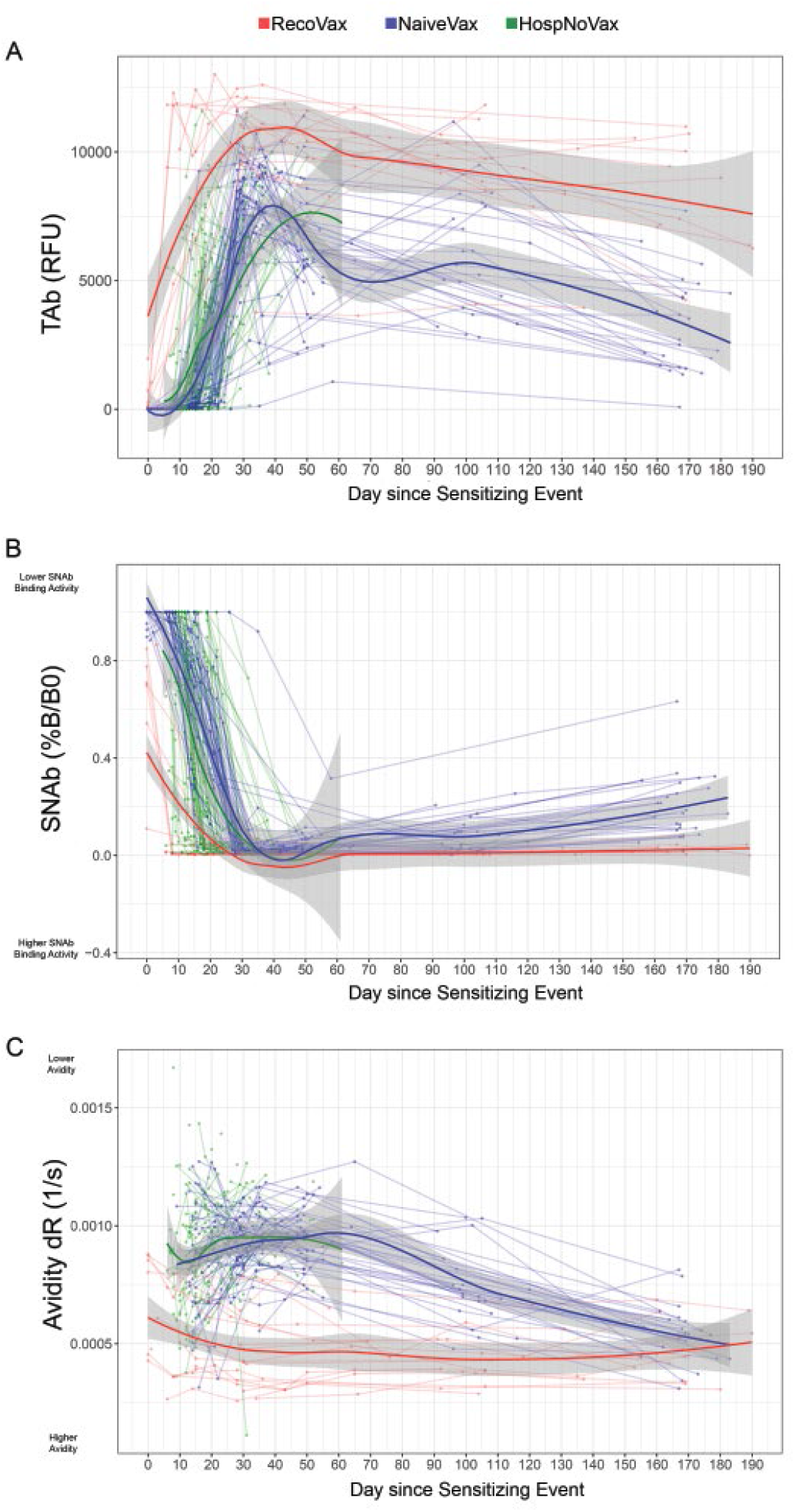
Dynamics of the anti-SARS-CoV-2 antibody response after vaccination or infection utilizing regression models. TAb (A), SNAb (B) levels and avidity (C) are displayed over time. A total of 686 data points were plotted from 19 RecoVax individuals (red), 49 NaïveVax individuals (blue) and 122 HospNoVax patients (green). All participants received the second dose 21 days after the 1^st^ dose. The trend of antibody level overtime was described by applying Muggeo’s method of estimating regression models with unknown break-points to estimate the changing time points of the trends.

NaïveVax displayed a slight lag in responding after the first dose of the vaccine in comparison to RecoVax, but increased at a similar rate to that of HospNoVax post infection -at an rate of 280 RFU/day (p<0.001). At approximately day 34 post D1, TAb began its slow overall decline at 29.4 RFU/day until the end of the follow-up period of up to 183 days post-D1 (p<0.001). (Figure 2A)

This is in stark contrast to the robust TAb response within the first week of vaccination in RecoVax. The TAb increased 1553 RFU/day for 7 days (p<0.001) at which point the RFU levels began to plateau and then begin to wane at a slower rate than NaïveVax at 14.72 RFU/day (p<0.001). (Figure 2A)

To complement the regression model, the TAb, neutralizing activity and avidity levels in all three cohorts were analyzed by stratifying into three 2-week time periods: 0-13 days (0-2 weeks), 14-27 days (2-4 weeks), and 28-42 days (4-6 weeks) after infection or D1. Data is provided in Supplemental Figure 1 in the Supplement. Additional comparisons were made between the vaccinated cohorts and OutPtNoVax at 4-6 weeks and 6-8 weeks post infection or D1. Data is provided in Supplemental Figure 2 in the Supplement. Figure 4 displays the antibody response at 4-6 weeks post infection or D1, a time period that was available for comparison in all four cohorts in the retrospective and prospective arms of the study.

Of note, RecoVax consistently had higher TAb levels than NaïveVax, HospNoVax and OutPtNoVax during the first 1.5 – 2 months post vaccination or infection (supplemental Figures 1A and 2A; Figure 4A). The RecoVax TAb was substantially increased in the first two weeks post-D1 with a median TAb of 882 RFU (IQR 93-9916) and continued to rise to a median TAb of 9916 (IQR 7455-9916; p=0.001)) during weeks 2-4. This remained stable during this 2 month follow-up period comparison, with a median TAb of 9916 (IQR 9660-9916) and 9916 (IQR 9095-9916) at 4-6 and 6-8 weeks, respectively (p=0.801).

Although initially NaïveVax TAb remained lower up to 4 weeks post D1 in comparison to HospNoVax (TAb of 430; IQR 208-1241 versus 3414; IQR311-5843; p<0.001) the TAb was significantly higher after D2. (Supplemental Figure 1A) At 4-6 week after D1, the NaïveVax TAb was 1.2 fold higher than HospNoVax and 7.7 fold higher than OutPtNoVax. (NaïveVax TAb: 7919 [IQR 6832 -9339]; HospNoVax TAb 6683 [IQR5573-8490]; OutPtNoVax Tab 1029 [164-2966]; p<0.001). (Figure 4, Supplemental Figures 1A and 2A).

### Qualitative antibody response during first 2 month in vaccinated versus SARS-CoV-2 infected individuals

#### Neutralization Antibody Levels by SNAb

In all three cohorts, the neutralization activity gradually increased over time, albeit at different time scales (Figure 2B, Supplemental Figure 1B; note: neutralizing activity is inversely proportional to the %B/B0 in the figures). In the regression models, HospNoVax had a gradual increase in neutralizing activity up to 24 days post-infection with a change in SNAb of 5.537 %B/B0 per day (p<0.001). The neutralization activity then began to wane but the change was insignificant (mean 7.716%B/B0; coefficient= -0.51442; p=0.055)

In comparison, NaïveVax displayed slight lag generating neutralizing activity after D1 with no changes from day 0-7 post-D1 (coefficient 0.1476; p=0.36). From days 7-30 post-D1, the neutralizing activity increased at a rate of 4.304 %B/B0 per day (p<0.001). Then, as seen with the TAB, there was loss of neutralizing activity after day 30 (p<0.001) at a rate of 0.115%B/B0 per day.

In comparison, RecoVax had a much more dramatic increase in neutralization activity. (Figure 2B) The neutralization activity improved at a rate of 9.381 %B/B0 in the first 8 days post-D1 (p=0.003), at which point the neutralization activity mildly began to wane at 0.008 %B/B0 per day (p=0.005).

As with the TAb, RecoVax displayed consistently higher neutralizing activity across all time periods when compared to the other cohorts. (Figure 4B, Supplemental Figure 1B and 2B). In the first 2 weeks alone, the SNAb was 8.0 and 6.4-fold better than that of NaïveVax and HospNoVax, respectively (p<0.001) (median SNAb %B/B0 were: RecoVax 12.48 IQR1.14-69.84; NaïveVax 100 IQR 95.53-100; HospNoVax 80.07 IQR 31.77-100). After the second dose of the vaccine, at 4-6 weeks post-infection/vaccination, the RecoVax SNAb neutralizing activity continued to remain higher than that of the NaïveVax, HospNoVax and OutPtNoVax cohorts (p<0.001). (Supplemental Figures 1B,2B and Figure 4)

Of note, NaïveVax SNAb at this 4-6 week time point had improved significantly and was at the levels found in HospNoVax (SNAb %B/B0 1.85 IQR 0.79-2.98 versus 1.19 IQR 0.58-4.22, respectively. p=0.367). Furthermore, the NaïveVax SNAb neutralizing activity was 14.8-fold higher than the OutPtNoVax (SNAb %B/B0 NaïveVax 1.85 IQR 0.79-2.98 versus OutPtNoVax 27.55 IQR 8.44-63.48; p <0.001) and remained 8.6-fold higher during the 6-8 week post vaccination period (SNAb %B/B0 NaïveVax 3.38 IQR 1.87-6.60 versus OutPtNoVax 29.03 IQR 9.05-55.25; p <0.001) (Supplemental Figures 1B and 2B; Figure 4B)

#### Antibody avidity

Generally, over the initial 6-week time period, the strengthening of antibody avidity in any of the three cohorts was a gradual process, as would be expected. (Figure 2C; Supplemental Figures 1C and 2C; note: avidity is inversely proportional to the relative dissociation rate [dR] in the figures). Using regression models, it was noted that the HospNoVax cohort avidity did not significantly change over time during its 61 day follow up period. (mean 8.998×10^−4^/s; coefficient =0.0208×10^−4^; p=0.162).

Likely due to the early production and then disappearance of IgM antibodies, NaïveVax appeared to display a period of avidity worsening with a dR change of 3.422×10^−6^/s per day (p=0.02). However at about 46 days post vaccination, avidity improved at a rate of 3.523×10^−6^/s per day (p<0.001)

RecoVax consistently held a stronger avidity in comparison to the other cohorts (Supplemental Figures 1C and 2C, Figure 4C). Its avidity slowly but consistently improved at a rate of 0.5×10^−6^/s per day. Its consistently stronger avidity was noted at multiple time points. For example, at 4-6 weeks post infection or D1 (Figure 4C), the median RecoVax avidity (median dR 4.24×10^−4^/s, IQR 4.323-5.195 ×10^−4^/s) was nearly 2.5-fold higher (p<0.001) than that of NaïveVax (4.24×10^−4^/s, IQR 4.323-5.195 ×10^−4^/s), HospNoVax (4.24×10^−4^/s, IQR 4.323-5.195 ×10^−4^/s) and OutPtNoVax (4.24×10^−4^/s, IQR 4.323-5.195 ×10^−4^/s). However, it should also be noted that the NaïveVax, HospNoVax and OutPtNoVax held a similar avidity at all time points (e.g. p=0.624 at 4-6 weeks).

### Comparison of antibody dynamics post-D1 and -D2 in RecoVax and NaïveVax upto ∼6 months post-vaccination

Comparisons were made between the two vaccinated cohorts by analyzing the antibody levels, SNAb and antibody avidity during key time periods in relation to the vaccine doses. (Figure 3) The data were binned into 1) a baseline time period [median 1 day; IQR 0-6 post-D1], 2) prior to D2 [median 16 days post-D1; IQR 15-21], 3) ∼2 weeks post-D2 [median 35 days post-D1; IQR 34-36], 4) ∼4 weeks post-D2 [median 49 days post-D1;IQR 49-52], 5) ∼3 months post-D1 [median 100 days post-D1; IQR 97-105] and 6) ∼6 months post D1 [median 168 days post D1;IQR 164-170].

**Figure 3.**
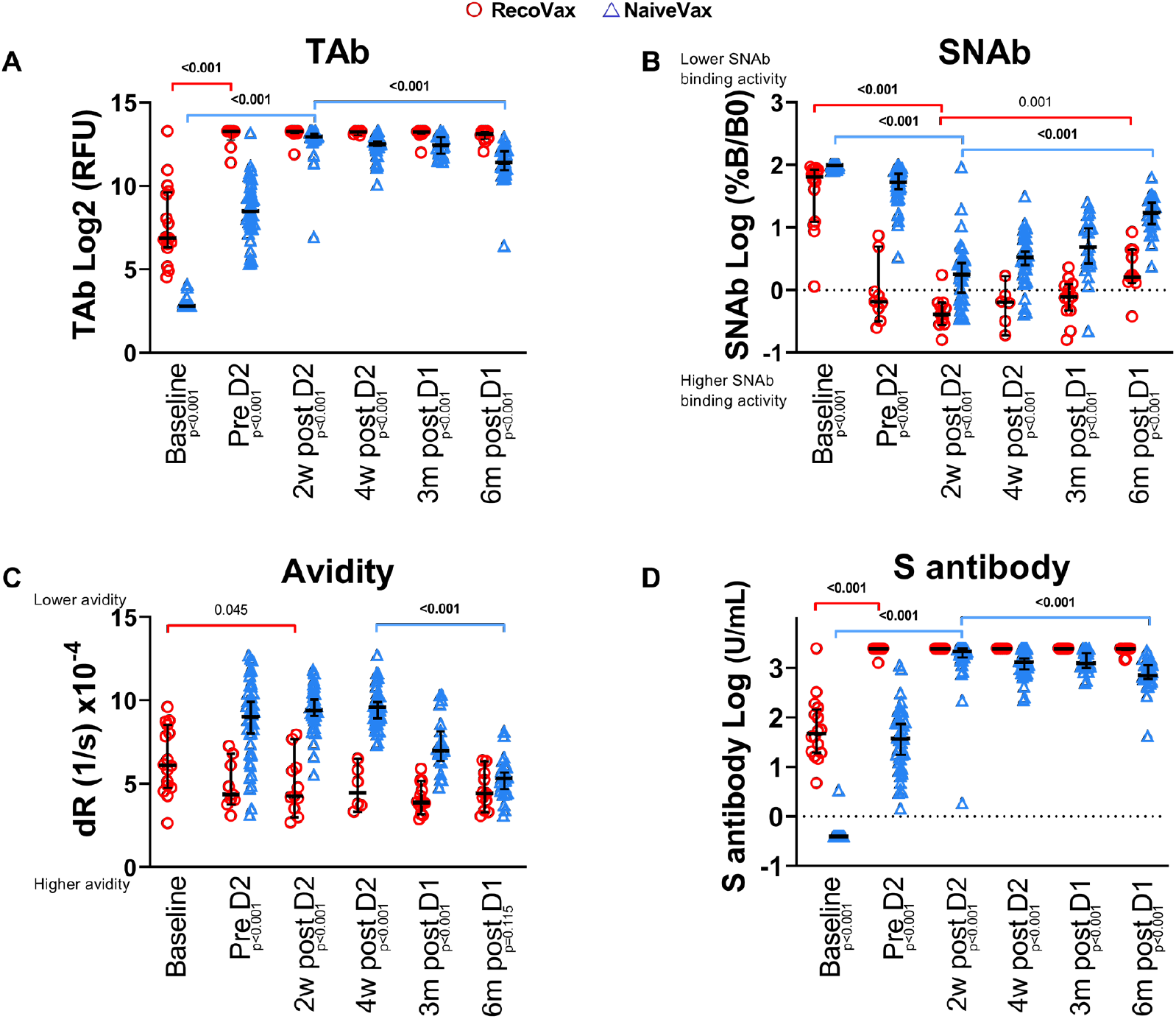
TAb (A) antibody response, SNAb (B) levels, avidity (C) and Anti-S (D) after RecoVax individuals (red circle), NaïveVax individuals (blue triangle) after first (D1) and second (D2) doses of the vaccine. Comparisons were made at baseline (median 1 day; IQR 0-6days post D1), prior to D2 (median 16 days post D1; IQR 15-21), ∼2 weeks post D2 (median 35 days post D1; IQR 34-36), ∼4 weeks post D2 (median 49 days post D1;IQR 49-52), ∼3 months post D1 (median 100 days post D1; IQR 97-105) and ∼6 months post D1 (median 168 days post D1;IQR 164-170) Horizontal black lines represent median values and whiskers represent 95% CI. Wilcoxon rank-sum was used for paired comparison while Kruskal-Wallis test was used for the comparison of three or more groups.

**Figure 4.**
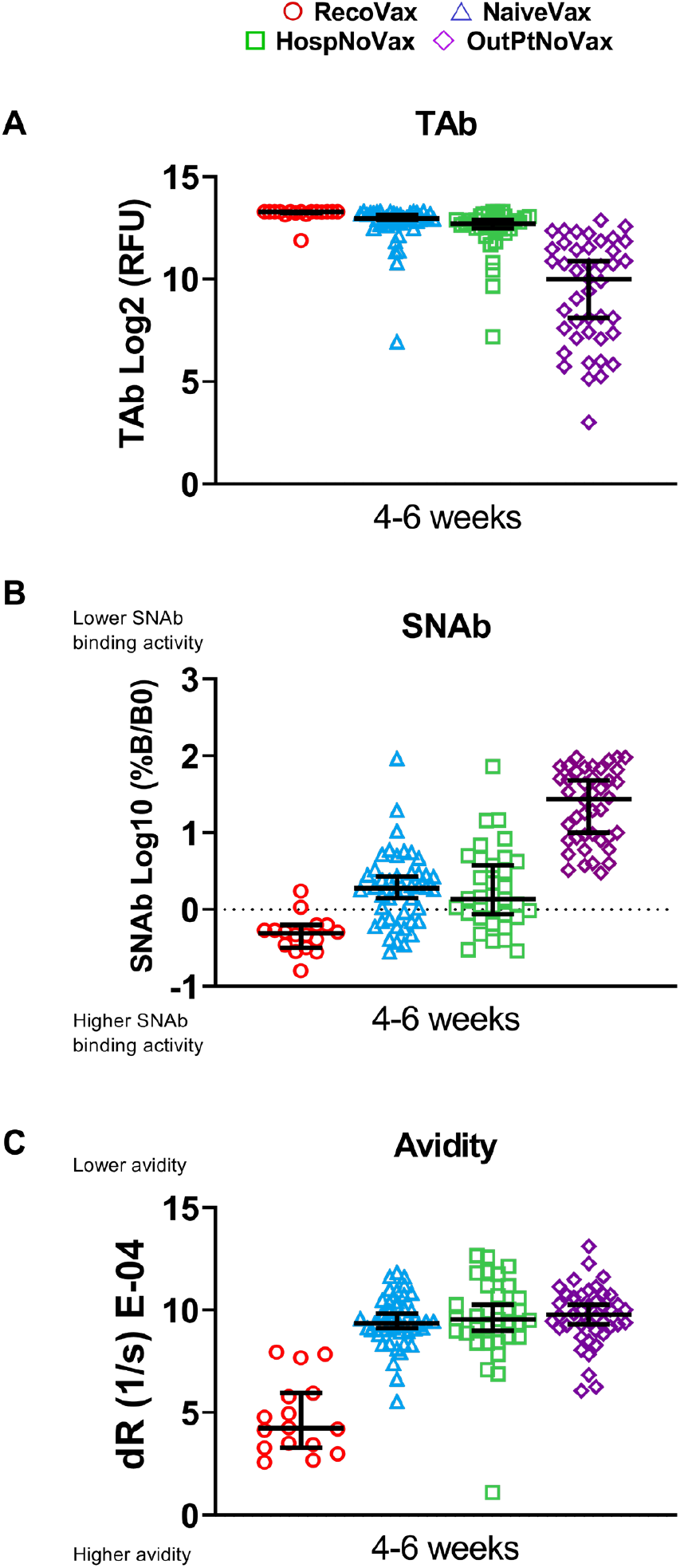
TAb (A) antibody response, SNAb (B) levels and avidity (C) after 4-6 weeks after vaccination or infection in RecoVax individuals (red circle), NaïveVax individuals (blue triangle), HospNoVax (green square) and OutPtNoVax (violet diamond). Horizontal black lines represent median values and whiskers represent 95% CI. Wilcoxon rank-sum was used for paired comparison while Kruskal-Wallis test was used for the comparison of three or more groups.

#### Quantitative antibody changes over time

RecoVax had TAb already present at their baseline (median 118 RFU; IQR 81.25-792 RFU) and these levels rapidly increased post-D1 (9916 RFU; IQR 7154-9916; p<0.001), prior to D2. Although there was a mild decrease in TAb at the ∼6 month time point (8997 RFU; IQR 7179-9916), this was statistically insignificant (p=0.698) and the levels remained relatively unchanged throughout the study period.

This is in stark contrast to NaïveVax’s TAb, which had a more gradual increase in TAb levels, with most individuals (40/41; 97.6%) not displaying a positive TAb during the first week post-D1. But all NaïveVax individuals did mount an antibody response in the following weeks prior to receiving D2, with a median TAb of 364.5 RFU (IQR 205.3-782.3). However, this was still 27-fold lower than the median TAb in RecoVax. (p<0.001). Maximal NaïveVax TAb levels were not achieved until 2 weeks post-D2 (median 7919 RFU; IQR 7253-9170) but this TAb decreased by over 50% overtime, with a median TAB 2706 RFU (IQR 1667-4511; p<0.001) at ∼6 months post-D1 and were 3.3-fold lower compared to RecoVax (p<0.001) at this time point. (Figure 3A)

As a comparison sub-study to an EUA platform, these results were confirmed by Elecsys Anti-SARS-CoV-2-S-antigen assay and similar patterns in the antibody response post-vaccination were observed. RecoVax had detectable levels of the anti-S total antibody prior to vaccination and following D1 increased from a median baseline of 47.45 U/mL (IQR 19.59-148.7) to above the upper limit of detection (>2500 U/mL). The RecoVax cohort median remained at this level at ∼6 months post-D1; however three individuals had levels below 2500 U/mL (median 2500 U/mL (IQR 2400 to >2500). In contrast, NaïveVax antibody levels gradually increased over time, with a median level of 37.77 U/mL (IQR 12.93-80.45 U/mL) prior to D2. These levels were boosted after D2 to a median of 2177 U/mL (IQR 1605 ->2500 U/mL) 2 weeks post-D2 (p<0.001). However, levels decreased to a median of 720U/mL (IQR 565-1269; p<0.001) at ∼6 months post-D2. Despite a robust increase after D2, the anti-S total antibody levels consistently remained lower than those of RecoVax (p<0.001 at all time points) (Figure 3D).

#### Neutralization Antibody changes over time in vaccinated subjects

In comparing the SNAb levels in NaïveVax to RecoVax, RecoVax displayed neutralizing capabilities at baseline, with a median SNAb of 65.42 %B/B0 (IQR19.47-84.59), which is 1.5-fold greater than NaïveVax (100%B/B0; IQR 96.09-100; p<0.001). As described previously, the neutralization capability seen in RecoVax improved dramatically after D1, prior to D2 (0.6600 %B/B0; IQR 0.4150-2.955; p<0.001). Although neutralizing capability remained unchanged through the ∼3 month period (0.7900 %B/B0; IQR 0.4741-1.217; p=0.144), at ∼6month post D1, neutralizing capability began to wane (1.610 %B/B0; IQR1.359-4.424; p=0.002).

Neutralizing activity was 81-fold lower in NaïveVax prior to administration of the second vaccine dose (median 53.57 %B/B0; IQR 31.29-77.95; p<0.001). Although the neutralizing activity of NaïveVax also improved with time, it remained 5.2-fold lower than RecoVax at 4 weeks post-D2 (3.380%B/B0; IQR 1.8015-6.603; p<0.001), 6.1-fold lower ∼3 months post-D1 (4.867%B/B0; IQR 2.544-11.17; p<0.001) and 10.8-fold lower ∼6 months post-D1 (17.35%B/B0; IQR 10.81-28.76; p<0.001). Also of note, neutralization activity began to wane in NaïveVax at 4 weeks post D2 and continued to decrease at the ∼6month time point (p<0.001). (Figure 3B)

#### Antibody avidity changes over time in vaccinated subjects

When comparing the avidity levels between RecoVax and NaïveVax cohorts, RecoVax had higher avidity levels prior to D2, twice that of NaïveVax (dR:4.372/s [IQR 3.904-6.465] versus 9.018/s [IQR 6.758-10.44] ×10^−4^/s; p<0.001). This gap in avidity levels between the two cohorts was observed over all time periods upto ∼3months. At 4 weeks post-D2, RecoVax still maintained a median avidity twice that of NaïveVax (dR:4.463/s [IQR 3.623-6.000] versus 9.605/s [IQR 8.773-10.29]×10^−4^/s; p<0.001) and 1.8-fold at 3 months post-vaccination (dR:3.889 [IQR 3.464-4.890] versus 7.000 [IQR 6.335-8.380] ×10^−4^/s; p<0.001). However, at ∼6months post D1, the gap in avidity closed with RecoVax avidity at 4.432 (IQR 3.390-5.642) ×10^−4^/s and NaïveVax avidity at 5.362 (IQR 4.509-5.977) ×10^−4^/s (p=0.115). (Figure 3C)

## Discussion

The following conclusions can be drawn from our study. First, RecoVax individuals exhibited a rapid anamnestic SARS-CoV-2 TAb and anti-S antibody response within days after D1 and these levels persisted up to the ∼6-month post-vaccination follow-up period. Second, the antibody response did not further increase after the second vaccine dose in this population. This is in contrast to the other cohorts, where TAb levels never fully matched RecoVax and in some cases decreased. Third, neutralizing activity was present at baseline in RecoVax and remained significantly higher in RecoVax versus the other cohorts, reaching a maximal plateau after D1 with no significant change until the ∼6-month follow-up period. However, at this point RecoVax SNAb began to wane (p=0.002). (Figures 2-3B) Finally, RecoVax had twice the avidity of the other cohorts in the first two weeks after the immunizing event and sustained this level of avidity throughout the study follow-up period. Nonetheless, NaïveVax’s continued avidity maturation achieved a similar avidity level by ∼6 months post-D1. Overall, the results of our study build upon and contribute to a growing body of evidence that vaccination generates similar, if not better, antibody levels to natural SARS-CoV-2 infection.(9, 12, 15, 16, 26)

To our knowledge, this is the first longitudinal study to compare avidity in these four included study populations. Antibodies with low intrinsic avidity are initially produced in the early humoral immune response and require time to mature and strengthen. Thus, it was not unexpected to find that NaïveVax, OutPtNoVax and HospNoVax generated antibodies with similarly lower avidity than those of RecoVax. Although it was expected that RecoVax at baseline had significantly higher antibody avidity (Figures 2-3C), it was notable that the antibodies induced by the vaccine showed robust avidity and the rate of avidity maturation was greater in NaïveVax in comparison to RecoVax (ΔdR 0.03523 versus 0.005645 ×10^−4^/s per day), allowing NaïveVax avidity to match that of RecoVax at ∼6months post D1. (Figure 2C and 3C) This observation, together with the knowledge that avidity continues to mature overtime (23), warrants additional investigation on whether long term avidity plays a clinically significant role in protection against SARS-CoV-2.

The immunological correlation of protection or thresholds required for vaccine efficacy against SARS-CoV-2 infection are not well defined. (27) Although not clinically recommended by the FDA(28), studies do attempt to correlate antibody levels to protection against the virus or disease when monitoring the immune response to vaccination, as antibody assays are generally convenient to perform. However, these may be non-mechanistic correlations and by no means absolute correlates. (29) Recently, studies (30, 31) have begun to define the relationship between SARS-COV-2 vaccine efficacy and neutralizing antibody titers by demonstrating significant correlation between vaccine efficacy and vaccine-induced neutralizing antibody activity. Neutralizing antibodies bind to viral targets and block entry into the cell, preventing infection of the cell with the virus. In this light, antibody avidity should not be discounted, as it is the binding strength of an antibody-antigen complex and could define the quality of the immune response.(32) Although avidity is typically low immediately after infection or vaccination, it undergoes maturation over a period of months and could reflect a better functioning active antibody pool. Therefore, future studies should be attempted in correlating antibody avidity with vaccine efficacy as antibody avidity may be a reliable and stable long term surrogate marker of immunological memory for infection. (33)

Such studies could also address concerns about the persistence and protective strength of the antibodies generated both by vaccination and natural infection. (34-36) There may be concern over antibody levels in those with mild COVID-19 symptoms (i.e. OutPtNoVax) as this population has overall lower antibody levels in comparison to individuals with more severe symptoms (37) (i.e. HospNoVax) or those that been vaccinated (Figure 4). Unfortunately, this study did not have adequate long-term data for avidity regression modeling in the OutPtNoVax cohort but early studies do reveal that avidity matures up to 1 year post infection.(26) Together with the proper future correlative studies, it may become reassuring that avidity in the OutPtNoVax, HospNoVax and NaïveVax have similar avidity (Figure 4C) and may continue to further strengthen over time, as demonstrated in NaïveVax, when avidity reached equivalent levels to that of RecoVax by ∼6months post D1 (Figure 3C).

Similar concerns over poor antibody production and protection against SARS-CoV-2 have arisen in at risk populations, such as those with hematological malignancies or transplant patients (38-40). Although these at risk individuals mount a poorer quantitative antibody response, it may be of interest to study their antibody avidity maturation profile and determine whether it could offer any protection against the virus.

### Limitations

Our study has several limitations. First, selection bias may exist in both the prospective and retrospective studies. As the HospNoVax and OutPtNoVax specimen in the study were retrospectively collected, the study was reliant on pre-existing data and remnant specimens. During this early time period of the pandemic in New York City, most hospitalized individuals were older, with more severe symptoms and predominately male,(41)which was reflected in our cohort (median age 67.5 years, IQR 54.0-77.0; 66% Male). The prospective study participants were younger, approaching or at middle age (overall median age 44.4 years, IQR 33.6-57.4) and predominantly female (74%). Indeed, the better population for comparison are OutPtNoVax, but there was insufficient data to perform a direct comparison at those similar timepoints

Additional bias may exist with the prospective vaccination study volunteers, as study participants were healthcare workers (eligible for the vaccine in late December 2020 to January 2021), possibly reflecting a study population with fewer comorbidities. Together with the small sample size (n=60), these results may not represent those of the general population.(42, 43)

Second, given the exploratory nature of the study and limited sample size, post-hoc adjustments were not performed. This small study size also prevented multivariate analysis to look for possible confounders between the cohorts. For example, a previous study found SNAb to be age associated in SARS-CoV-2 infected individuals.(42) The association of age with TAb, SNAb and avidity was explored in this study, but no association was found, likely due to the predominately middle-aged cohort in this study.

Thirdly, the time period between symptom onset and antibody testing in HospNoVax is arguably not an exact time equivalent to the time period of antibody testing post-vaccination as the incubation period and symptom onset post-infection can be up to 14 days.(24) However, it has been reported that the median incubation period is 5.1 days (25) and this was added to the OutPtNoVax and HospNoVax’s time since symptom onset in an attempt to overcome lead time bias. Additionally, the follow-up time for this cohort was limited compared to the vaccinated cohorts.

Finally, this study focused on the humoral response to the SARS-CoV-2 vaccine, as vaccine development strategies are designed to maximize this response. However, the T-cell compartment also plays a major role in the immune response(44). SARS-CoV-2 infection has been shown to induce the T-cell immune response, which play a major role in preventing severe disease.(45, 46) Although this is beyond the scope of this study the interplay between the T-cellular response and humoral response may also provide important insights into better correlative studies for vaccine effectiveness.

## Conclusions

Our data suggest that two doses of the mRNA vaccine are warranted in NaïveVax individuals to achieve a similar early antibody response to RecoVax individuals. Individuals with mild COVID-19 symptoms (OutPtNoVax) overall maintained lower antibody levels compared to the vaccinated cohorts, especially warranting vaccination despite prior infection. Furthermore, as the vaccine elicited a maximal antibody response after only one vaccine dose in RecoVax, one dose may be sufficient in this population. Although longer term longitudinal studies are required, the persistent TAb and avidity in this population may support a single dose vaccination strategy. This would be a resource-conscious solution to help address global vaccine shortages. Monitoring individuals for antibody titers long term (as is done with the Hepatitis B or MMR vaccines), as well as monitoring neutralizing activity and avidity(23), may be prudent in determining the vaccine efficacy and the need for future booster vaccinations.

## Materials and Methods

### Sources of serum specimens and data acquisition

The standard practice for serum collection and storage in the clinical laboratories involves collecting venous blood into a serum separator tube and centrifuging the specimen (1500g for 7 minutes) within 2 hours of collection to separate cells from serum. Patient and study participant serum samples after routine clinical testing were stored at -80ºC until further analysis was performed.

#### Retrospective study of COVID-19 outpatients and hospitalized patients

The retrospective study included a cohort of 122 adult patients who presented to the ED and were subsequently hospitalized at NYP/WCMC from 3/8/2020 to 4/7/2020. This cohort was described in a previous assay validation study.(23) All patients in this cohort tested positive for SARS-CoV-2 by real-time reverse transcription polymerase chain reaction (RT-PCR) within one day of the ED visit. In total, 317 remnant serum samples were collected and frozen during the hospitalization for future analysis.

Another cohort from this retrospective study included 160 convalescent COVID-19 patients who were seropositive in the outpatient setting at NYP/WCMC from 4/30/2020 to 5/20/2020. All participants in this study were adult, non-pregnant patients who were not hospitalized (previously or at the time of antibody testing) due to SARS-CoV-2 infection. In total, 160 remnant serum samples were collected and frozen for this future analysis.

Demographic data and date of symptom onset was collected from the electronic medical record for both cohorts (Allscripts, Chicago, IL).

#### Prospective study of vaccinated individuals

Sixty-one healthcare workers at NYP/WCMC vaccinated with two doses of the BNT162b2 SARS-CoV-2 mRNA vaccine (Pfizer, New York, NY) were included in this study. (Figure1) Participants were asked to donate blood samples for serologic analysis within a week of the 1^st^ dose of the vaccine (D1), approximately 2 weeks and 4 weeks after each vaccine dose, and approximately 3 and 6 months after D1. Although not all of the 61 participants provided samples at all time points, all specimens were considered for analysis in this study, as indicated in the figures and tables. A total of 326 serum samples were prospectively collected 12/31/2020-7/1/2021. (Figure 1)

An additional 7 participants from the NYP-WELCOME study (47) were also included in the prospective aspect of this study. Forty-three specimens had been collected from this cohort during the time period 12/11/2020-7/6/2021 and frozen in the Weill Cornell Medicine BioBank for future analysis. (Figure 1)

### SARS-CoV-2 total RBD antibody, surrogate neutralizing antibody assay and avidity assays

The SARS-CoV-2 total receptor-binding domain (RBD) antibody (TAb), surrogate neutralizing antibody (SNAb) and avidity were used to measure serum antibody levels on the TOP-Plus (Pylon 3D analyzer; ET HealthCare, Palo Alto, CA), and were previously described. (23)

The TAb assay measures the overall interaction between SARS-CoV-2 antibodies and the RBD of the virus spike (S) protein, with a read out of sample relative fluorescence unit (RFU).

SNAb is a competitive binding assay, based on the anti-SARS-CoV-2 antibody-mediated inhibition of the interaction between the ACE2 receptor protein and the RBD. The assay readout is the percentage of RBD-ACE2 binding [%B/B0; (sample RFU/negative control RFU) *100%], which is inversely associated with neutralizing activity. It was previously shown (23) to correlate well with two established SARS-CoV-2 virus neutralization tests (plaque reduction neutralization test and pseudo virus neutralization test) and was used in this study to evaluate the neutralization activity of the antibodies generated post-vaccination and post-infection in ED COVID-19 patients in this study.

The avidity assay provides the calculated relative dissociation rate (dR; 1/s). This measurement is inversely associated with antibody avidity. A higher intrinsic binding strength of a paratope to RBD or addition of paratopes to the antibody structure results in a higher binding strength, which results in a lower dR of the antibody-RBD pair. The assay had good correlation with the Bio-Layer Interferometry avidity assay, another assay used for measuring antibody avidity.(23)

### The Roche Elecsys Anti-SARS-CoV-2S-and N-antigen assays

The Elecsys^®^Anti-SARS-CoV-2S and Anti-SARS-CoV-2 electrochemiluminescence immunoassay were used to detect antibodies to SARS-CoV-2 spike (S) RBD and the nucleocapsid (N) antigen, respectively, in the serum samples. These were performed on the Roche Cobas e411 (Roche Diagnostics, Indianapolis, IN). These assays received EUA approval from the United States Food and Drug Administration (FDA). The Anti-SARS-CoV-2S Elecsys^®^assay was used for comparison with the TOP-TAb assay. The Anti-SARS-CoV-2 assay was used in identifying or confirming previously infected individuals in the vaccinated cohort.

### Statistical analysis

The trend of antibody level overtime was described by applying Muggeo’s method of estimating regression models with unknown break-points to estimate the changing time points of the trends.(48) A linear mixed effect model was fitted for each segment of time based on estimated break-point to show the trend for the time period between break-points. As indicated, coefficients and p-values from regressions were reported. To visualize the trend, trajectories were plotted via a smooth line with loess method for each group. Only data up to day 61 post-infection were available for this analysis in HospNoVax and comparisons were only performed up to this time period.

Wilcoxon rank-sum and signed-rank tests were used between numerical variables and paired comparison, respectively. Kruskal-Wallis test was used for the comparison of three or more groups. Bivariate associations between outcome variables and clinical parameters were evaluated using Fisher’s exact test or chi-squared test, as appropriate. Descriptive data were presented as median with interquartile range (IQR) unless otherwise specified. p<0.05 were considered significant.

Analyses were performed in statistical programming language R version 4.0.2 (2020-06-22) or in GraphPad Prism Version 9.1.2 (GraphPad Software, La Jolla, CA).

### Study approval

The retrospective (IRB#20-03021671) and prospective (IRB#20-11022929; IRB#20-04021831) studies in this manuscript were performed at NewYork–Presbyterian Hospital/Weill Cornell Medical Center (NYP/WCMC) with approval by the Institutional Review Board of Weill Cornell Medicine (WCM).

## Data Availability

Any data inquires may be made to the corresponding authors and will be reviewed according the to NYP/Weill Cornell institutional guidelines.

## Author contribution

SRB for conceptualization, investigation, supervision of project, data analysis and writing the manuscript; JY for sample collection, performing serology experimentation and editing the manuscript; AS for sample collection and performing serology experimentation; YQ for statistical analysis and editing the manuscript; MK and RZ for the assay development and providing analytical data analysis; SR for data collection, data collection design and general administrative support; PB for recruitment efforts; YH for sample collection; HSY for conceptualization and editing the manuscript; FSA and QHM for conceptualization and editing the manuscript; YYS for conceptualization; AC for editing the manuscript; EG and SF for design and implementation of the NYP-WELCOME study and editing of the manuscript; MMC for supervision of project and editing the manuscript; ZZ for conceptualization, investigation, supervision of the project, data analysis, and editing the manuscript.

## Acknowledgement

This research was partially funded by a COVID-19 research grant from Weill Cornell Medicine (Cushing). The NYP-WELCOME study was supported by Weill Cornell Medicine Competitive Research Funding (Formenti). ET-Healthcare provided seed instruments and reagents (Zhao). Roche Diagnostics provided support via Investigator initiated research funding (Racine-Brzostek and Zhao).

The authors would like to acknowledge Courtney Johnson-Williams, Jennifer Lopez, William Rodriguez, Rosanna Tineo and Jasmine Brown for their assistance with phlebotomy services. We also appreciate the administrative assistance of Sandra Aranibar-Atristain and Rita Frenzel and Maria Salpietro for her assistance in her role as the Director of the Weill Cornell Institutional Biobank.

## Abbreviation

COVID-19: Coronavirus Disease-2019
D1: 1st dose vaccine
D2: 2nd dose vaccine
DAOS: days after onset of symptoms
dR: relative dissociation rate
ED: Emergency Department
EUA: emergency use authorization
FDA: U.S. Food and Drug Administration
HospNoVax: hospitalized acutely infected non-vaccinated cohort
NaïveVax: naive vaccinated cohort
N: nucleocapsid
NYP: New York Presbyterian
OutPtNoVax: convalescent non-vaccinated COVID-19 outpatients
PI: days post-infection (SARS-CoV-2)
PRNT: plaque reduction neutralization test
PsV: pseudovirus neutralization test
RBD: receptor-binding domain
RecoVax: recovered vaccinated cohort
RFU: relative fluorescence unit
RT-PCR: real-time reverse transcription polymerase chain reaction
SARS-CoV-2: severe acute respiratory syndrome coronavirus 2
SD: standard deviation
SNAB: surrogate neutralizing antibody
TAb: total antibody
WCMC: Weill Cornell Medical Center.

