## Supplemental Data for "More rapid, robust and sustainable antibody responses to mRNA COVID-19 vaccine in convalescent COVID-19 individuals"

### Supplemental Material

#### Supplemental Methods

*SARS-CoV-2 antibody avidity assay*

The avidity assay is part of the TOP-Plus (Pylon 3D analyzer; ET HealthCare, Palo Alto, CA) biosensor platform and was previously described in detail and validated. (1) Briefly, the assay measures the rate of SARS-CoV-2 specific antibodies dissociated from RBD. This measurement is inversely correlated with SARS-CoV-2 antibody avidity. With washes between each step, the RBD pre-coated probe is sequentially incubated in diluted serum sample, biotinylated RBD and then Streptavidin-Cy5 conjugate to detect SARS-CoV-2 antibodies. The fluorescent signal before dissociation is the measured (Signal_0) and then the probe with the immobilized immunocomplex enters into multiple repetitive dissociation cycles of incubation in PBST dissociation buffer (PBS with 0.05% Tween20, pH 7.4). The fluorescent signal is measured at the end of each interval (Signal_t). Dissociation of Covid-19 antibody from either the coated RBD on the probe surface or the biotinylated RBD (linked to Cy5-Streptavidin) could result in the separation of Cy5 from the probe surface and drop of signal. Fluorescent signal directly correlates to the amount of antibody bound on the probe at the time of measurement. The amount of antibody bound on the probe surface through dissociation relative to the total amount of antibody on the probe before dissociation ([bound]/[total]) is expressed by the ratio of fluorescent signal at each time point through dissociation over the fluorescent signal before dissociation starts (Signal_t / Signal_0). A dissociation profile is constructed by plotting the fluorescent signal ratio as a function of time. The relative dissociation rate (dR) is calculated by fitting the first order rate equation to the dissociation profile: Ln(Signal_t / Signal_0) =Ln([bound]/[total])=-dR t

#### Supplemental Figures

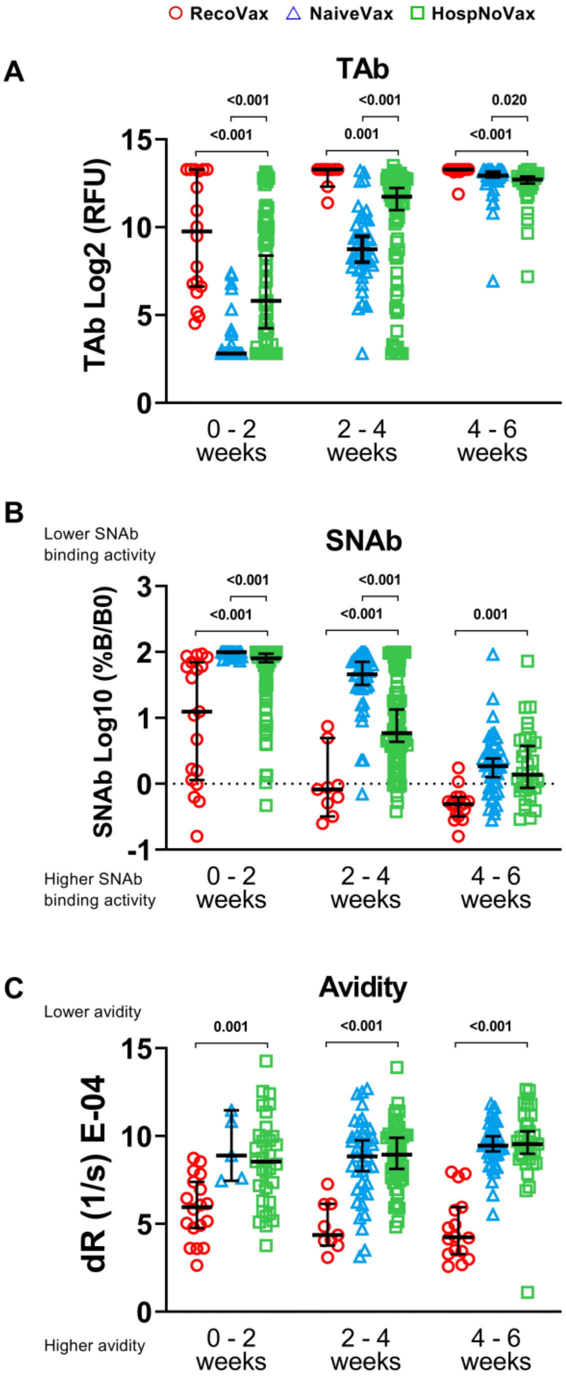


**Supplemental Figure 1:** TAb (A) antibody response, SNAb (B) levels and avidity (C) after vaccination in RecoVax individuals (red circle), NaïveVax individuals (blue triangle) and HospNoVax patients (green square). The three cohorts were compared at 0-2 weeks (0-13 days), 2-4 weeks (14-27 days) and 4-6 weeks (28-41 days) post 1st dose vaccination or post-infection. Long black lines represent median values and whiskers represent 95% CI. Wilcoxon rank-sum was used for paired comparison while Kruskal-Wallis test was used for the comparison of three or more groups.

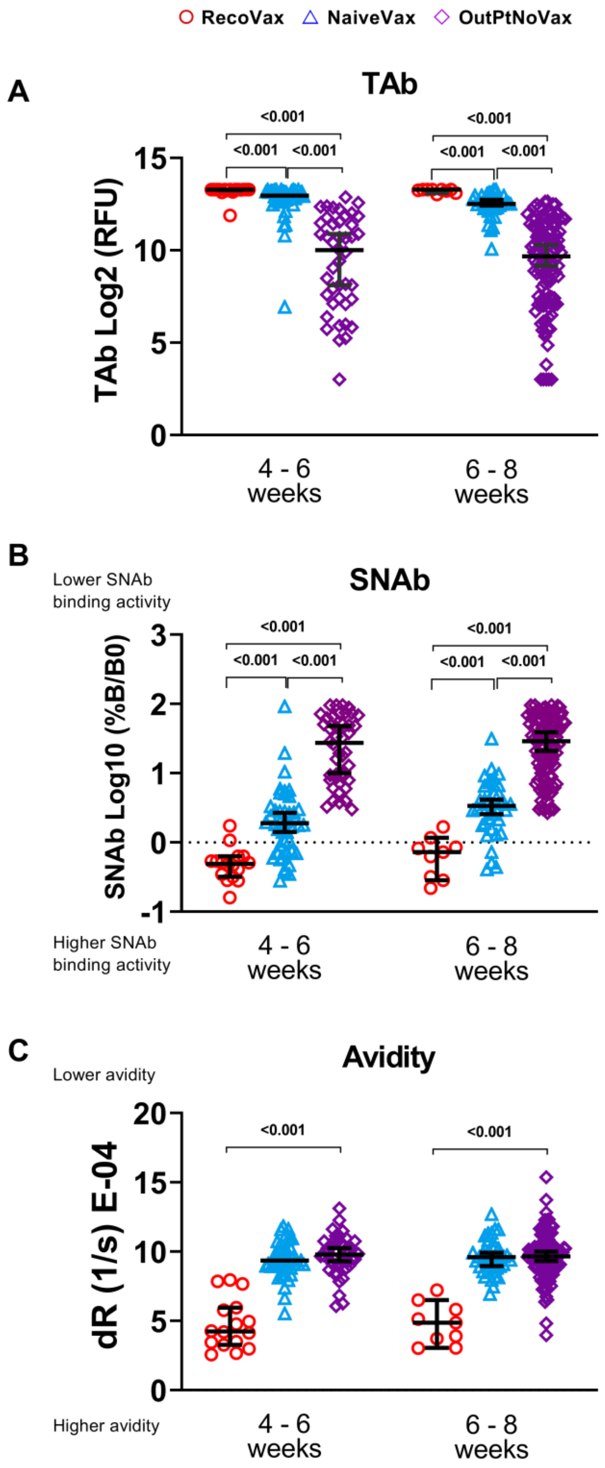


**Supplemental Figure 2:** TAb (A) antibody response, SNAb (B) levels and avidity (C) after vaccination in RecoVax individuals (red circle), NaïveVax individuals (blue triangle) and OutPtNoVax patients (violet diamond). The three cohorts were compared at 4-6 weeks (28-41 days) and 6-8 weeks (42-56 days) post- 1$^{st}$ dose vaccination or post-infection. Long black horizontal lines represent median values and whiskers represent 95% CI. Wilcoxon rank-sum was used for paired comparison while Kruskal-Wallis test was used for the comparison of three or more groups.
